# Identification of BcenGI15 genomic island harboring ST839 clone in a population of *Burkholderia cenocepacia* complex from a major tertiary care hospital in Northern India

**DOI:** 10.1101/2022.10.04.22280591

**Authors:** Tanu Saroha, Charu Singh, Sunil Kumar, Rajesh Kumar, Prashant P. Patil, Lipika Singhal, Vikas Gautam, Prabhu B. Patil

**Affiliations:** Bacterial Genomics and Evolution Laboratory, CSIR-Institute of Microbial Technology, Sector -39A, Chandigarh, India; Academy of Scientific and Innovative Research (AcSIR), Ghaziabad, India; Department of Biotechnology, Maharishi Markandeshwar (Deemed to be) University Mullana, Ambala, Haryana, India; Department of Microbiology, Institute of Medical Sciences, Banaras Hindu University Varanasi, India; Department of Microbiology, Government Medical College and Hospital, Sector -32B, Chandigarh, India; Dept. of Medical Microbiology, Post Graduate Institute of Medical Education and Research (PGIMER), Sector -12 Chandigarh, India

**Keywords:** *Burkholderia cepacia* complex, genomic island, antibiotic susceptibility, multilocus sequence typing

## Abstract

**Introduction:** *Burkholderia cepacia* complex (Bcc) is a non-fermenting Gram-negative bacilli (NFGNB) cluster with high genome plasticity and large genome size. As a major nosocomial pathogen, it is known to cause bacteremia, infections in cystic fibrosis patients. One of the factors contributing to multidrug resistance, virulence, and fitness is through chromosomally encoded genetic elements. They carry advantageous genes benefitting the host, thus its crucial to understand their stability and transfer in population. In an earlier study, we have reported a novel genomic island BcenGI15 in a unique clone of Bcc, ST824, involved in a major sepsis outbreak of a pediatric ward in an Indian hospital. In the present study, we have carried out screening of this genomic island by polymerase chain reaction (PCR) in an extensive collection of Bcc isolates from a major tertiary care hospital in Northern part of India.

**Materials and methods:** 90 isolates obtained from routine patient specimens over a period of 9 years revived from glycerol stock and identified as *Burkholderia cenocepacia* based on conventional biochemical tests, recA PCR-based restriction fragment length polymorphism (RFLP), and matrix-assisted laser desorption ionization time-of-flight mass spectrometry (MALDI-TOF MS). Isolates were screened for genomic island BcenGI15 via PCR using *attL* gene primers. Island positive isolates were subjected to multilocus sequence typing (MLST) and antibiotic susceptibility testing.

**Results:** The PCR in 16/90 (17.77%) isolates came positive for the presence of BcenGI15. Multi-locus sequence typing (MLST) revealed that all the positive isolates are clonal and belong to a dominant sequence type (ST) ST839.

**Conclusion:** MLST data analysis suggested presence of BcenGI15 in two different STs (ST824, ST839) from hospitals in north and west part of India. This suggests probable movement and selection for this element in Indian population of Bcc isolates.

## INTRODUCTION

*Burkholderia cepacia* complex (Bcc) comprises a group of non-fermenting Gram-negative bacilli (NFGNB) isolated from various sources and has gained importance recently because of its high infection rate in chronic cystic fibrosis patients (1). There have been reports of bacteremia in immunocompromised and immunocompetent hosts as well. Bcc is usually of great concern to clinicians because it has intrinsic resistance to first and second-generation cephalosporins, aminoglycosides, antipseudomonal penicillin, and polymyxins due to altered outer membrane permeability barrier and efflux pumps. Acquired resistance is through the production of beta□lactamases and other modifying enzymes. The various mechanisms of acquiring resistance have always been the curiosity of scientists and various probable mechanisms are being studied.

A major driving force in bacterial evolution is the horizontal transfer of mobile genetic elements that are chromosomal or plasmid-borne. These include genomic islands (GEI), plasmids, integrons, etc. (2, 3). The genomic islands encode machinery for self-excision, conjugative transfer, and integration and are known as Integrative and Conjugative Element (ICE) (4, 5). Each ICE consists of core genes, which encode functions essential for maintenance and their self-transmission, and cargo genes, which render them the phenotypes essential for their niche adaptation (6). These are propagated by horizontal gene transfer (HGT) and replicated or transferred as a part of genetic material during the cell division. They are also known as conjugative transposons and act as essential carriers for acquiring many antibiotic resistance-associated genes among bacteria (7). HGT has a role in shaping the bacterial genome by allowing the rapid acquisition of unique genes with adaptive functions that can have a tremendous impact on the adaptation and evolution of bacteria (7, 8).

There is another class of GEI, known as Integrative Mobilizable Element, which has autonomous machinery for both integration and excision, but lacks the machinery for conjugation or encodes very few conjugation modules (7). They are non-self-mobilizable but can be mobilized with the aid of conjugation machinery of helper elements like conjugative plasmids and other ICEs (9). GEIs have a significant role in the adaption and evolution of bacteria, which can be explained by the propagating antibiotic resistance-associated genes and virulence factors that lead to the genesis of superbugs. It also has a significant role in the dispersion of catabolic genes in the environmental symbiotic and commensal bacteria, thus forming unique and new metabolic pathways in these bacteria (10).

There are fourteen GEIs in Bcc, named BcenGI1 to BcenGI14, which have already been reported in the reference genome *B. cenocepacia* J2315, and in continuation of that, we reported a novel GEI in a unique clone of *B. cenocepacia* from a Mumbai Hospital (Western part of India) outbreak and named it as BcenGI15 (11). When expanded multi-locus sequence typing was performed on three clinical isolates and two rubber stopper isolates of amikacin vials, the novel sequence type (ST) 824 was obtained from all the isolates proving an epidemiological link between these isolates. It identified the unopened amikacin vial as the source of this outbreak. It is plausible that element is driving the rapid evolution of Bcc isolates in India, and therefore, we screened an extensive collection of Bcc isolates from a major tertiary care center of North India for distribution and prevalence of BcenGI15 genomic island. We were interested in knowing whether the element or the clone has moved to another geographic location within India. Considering heavy patient density and tropical climatic conditions with heavy antimicrobial usage, systematic studies on the movement of novel GEIs in nosocomial isolates are a dire necessity.

## MATERIALS AND METHODS

A total of 90 isolates included in our study, previously stocked for earlier phylogenetic analysis and efflux pumps expression, were again revived, and used for screening BcenGI15 by polymerase chain reaction (PCR) (12). These isolates were obtained from routine patient specimens (blood, endotracheal aspirate, fluid, and pus) received over a period of 9 years 2005 – 2013 (Supplementary Table1). The tertiary care center is the biggest in the region and caters to patients from many states from the Northern part of India.

Isolates were identified using biochemical tests such as triple sugar iron agar test, amino acids decarboxylation test, and lead acetate paper strip test for hydrogen sulfide detection (13). Further, we confirmed their identification with matrix-assisted laser desorption ionization time-of-flight mass spectrometry (MALDI-TOF MS) (Bruker Corporation Ltd) (14) and recA PCR-based restriction fragment length polymorphism (RFLP) by commercially available PCR kit purchased from Qiagen Q solution Inc. Canada as per protocol (15).

The isolates were subjected to microbial susceptibility testing by Kirby-Bauer disk diffusion test (DD) according to recent CLSI (Clinical Laboratory Standards Institute) guidelines. This test was done for co-trimoxazole (COT) (1.25 μg/23.75 μg), ceftazidime (CAZ) (30 μg), tetracycline (TET) (30 μg), levofloxacin (LVX) (5 μg), meropenem (MEM) (10 μg), and minocycline (MIN) (30 μg). Minimum inhibitory concentrations (MIC) was also determined by agar dilution as per CLSI guidelines against minocycline (sensitive, S≤4 and resistant R≥16 μg/ml), levofloxacin (S≤2 & R≥8 μg/ml), and ceftazidime (S≤8 & R≥32 μg/ml).

90 isolates of *B. cenocepacia* were subjected to PCR targeting genomic island BcenGI15 (16). PCR was done using *attL* gene primers (Table1) following the same protocol as mentioned in a previous study authored by Patil *et al*. (16). Plating of the isolates was done on Muller-Hinton agar incubated overnight at 37°C and genomic DNA was extracted from the colonies obtained the next day by using a commercial kit (Bacterial DNA Mini-Prep Kit) supplied by Zymo Research Corporation (Orange, CA, USA) which was used as a template. PCR was carried out using a PCR master mix (Sigma Aldrich). The steps were followed as per the kit manufacturer’s instructions.

**Table 1.**
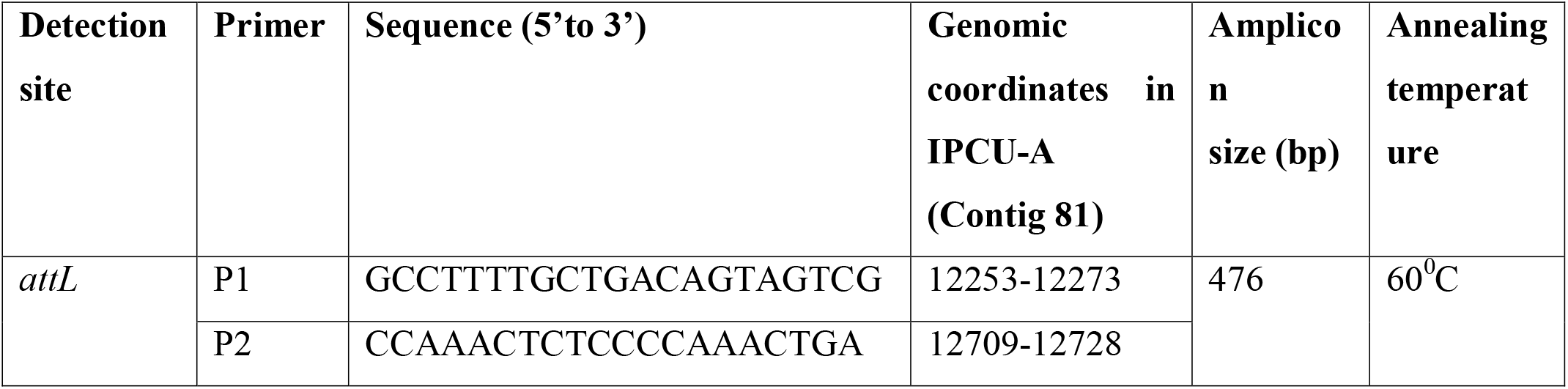
Primer Sequences of BcenGI15.

**Table 2.**
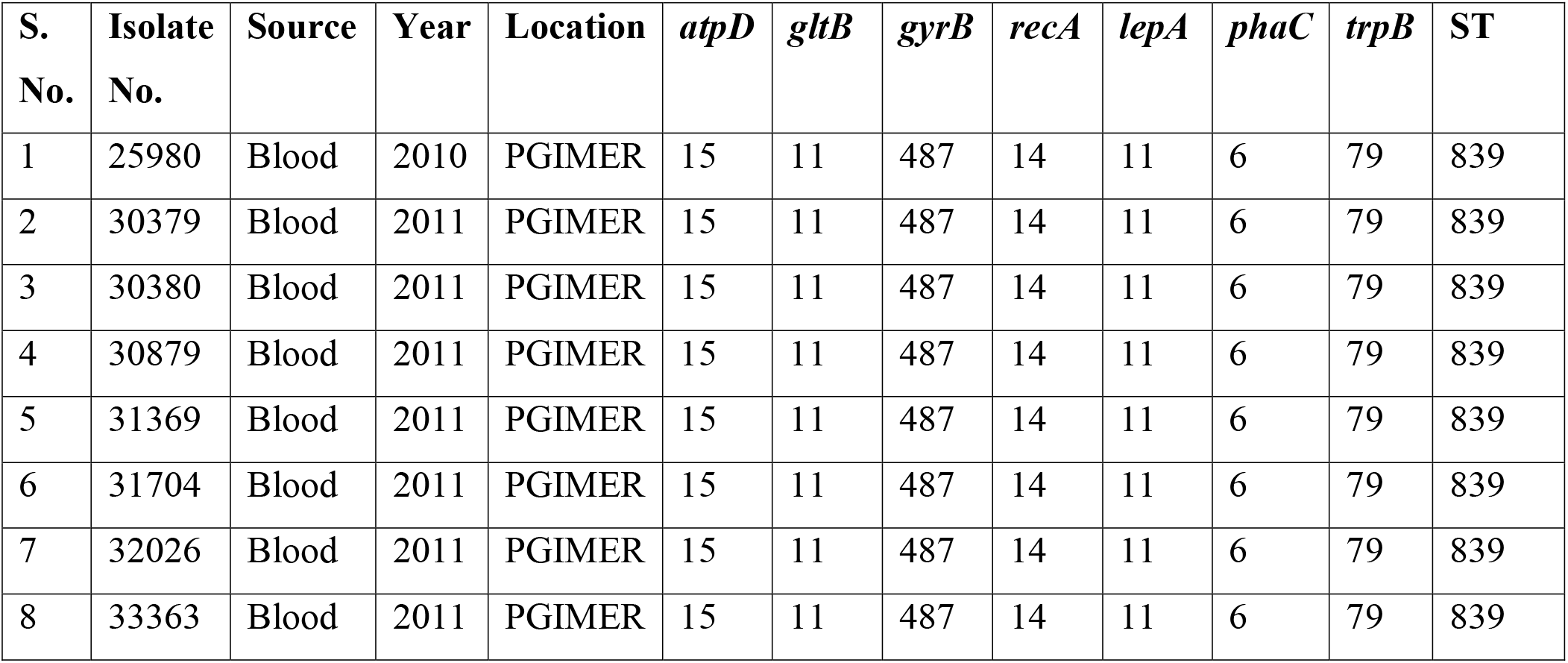
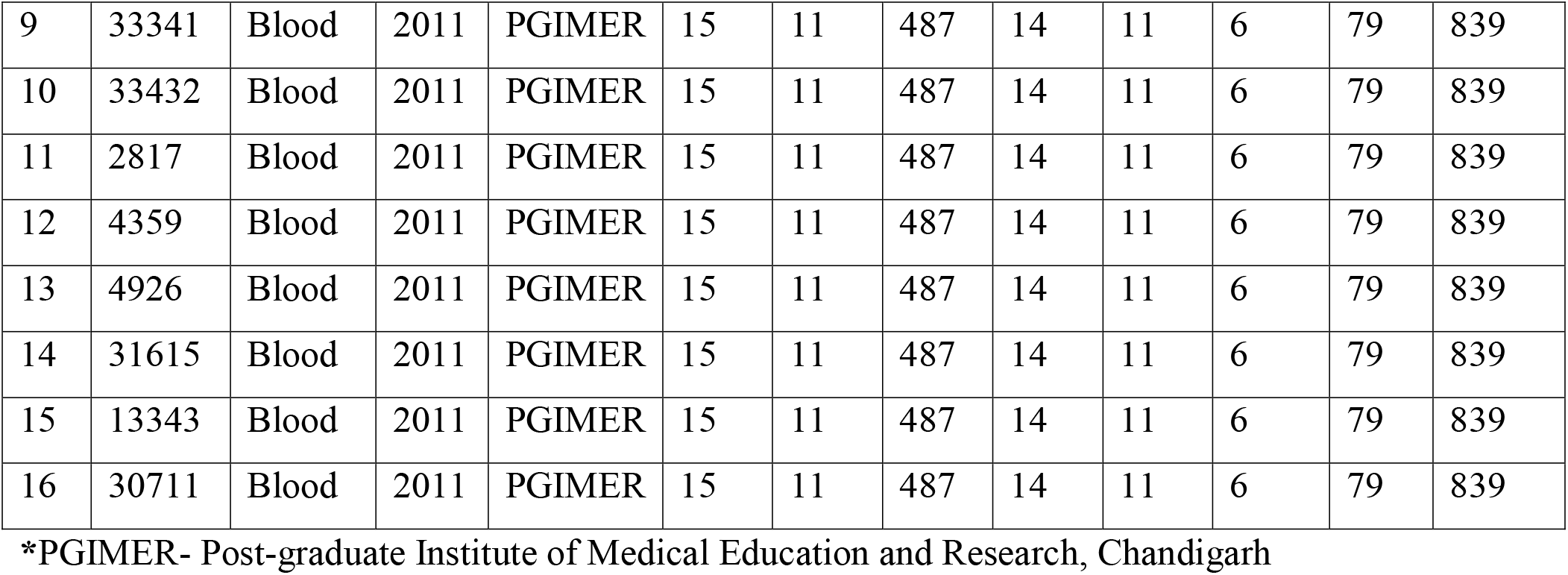
Sequence Types and allelic profile of Bcc isolates harboring mobile genetic element (BcenGI15) along with isolation details.

BcenGI15 positive isolates were subjected to Multi-locus sequence typing (MLST) by PCR amplification of seven house-keeping genes: ATP synthase beta chain (*atpD*), Glutamate synthase large subunit (*gltB*), DNA gyrase subunit B (*gyrB*), Recombinase A (*recA*), GTP binding protein (*lepA*), Acetoacetyl-CoA reductase (*phaC*), Tryptophan synthase subunit B (*trpB*) according to a previously published method (12). STs, allele profiles, and clonal complexes were assigned by the PubMLST database (www.pubMLST.org/bcc).

## RESULTS

Bcc isolates in this study were obtained from various specimens that included blood culture, body fluid, pus, endotracheal aspirate, and other respiratory specimens. Out of 90 *B. cenocepacia* isolates, 77(85.5%) were obtained from blood culture, 9(0.1%) from respiratory specimens, two isolates from endotracheal aspirate, and one each from fluid and pus (Supplementary Table1). These isolates were identified as NFGNB, using recA PCR-based RFLP and MALDI-TOF MS. The recA-PCR of isolates showing faint bands were repeated (15). All the isolates of Bcc were processed by PCR using *attL* gene primers of BcenGI15 (Table1). The PCR in 16/90 (17.77%) isolates came positive for the presence of the island. These isolates were from the year 2010 (1 isolate) and 2011(15 isolates). All isolates positive for BcenGI15 were obtained from blood culture. This indicates the presence of island BcenGI15 in isolates from another hospital located in the northern part of India, apart from the western part. It is important to note that in both the hospitals, the isolates from patients were associated with blood infection or bacteremia, or sepsis. While Mumbai isolates belonged to clone ST824, we were keen to know the clonality of the isolates isolated from a hospital in northern India.

Hence, we carried out MLST of all the sixteen isolates and it was interesting to note that all the isolates belonged to ST839. This suggests that the clone might have acquired BcenGI15 and expanded in the population. This indicates that probable movement of element in different STs in India.

Further, we have carried out antibiotic susceptibility test on island positive isolates (Table 3). All the isolates are resistant to tetracycline antibiotic though it is not a recommended drug for Bcc. Isolate 31704 is resistant to all antibiotics tested except for MIN and LVX. Almost all isolates are sensitive to CAZ, COT, MEM, and LVX.

**Table 3.**
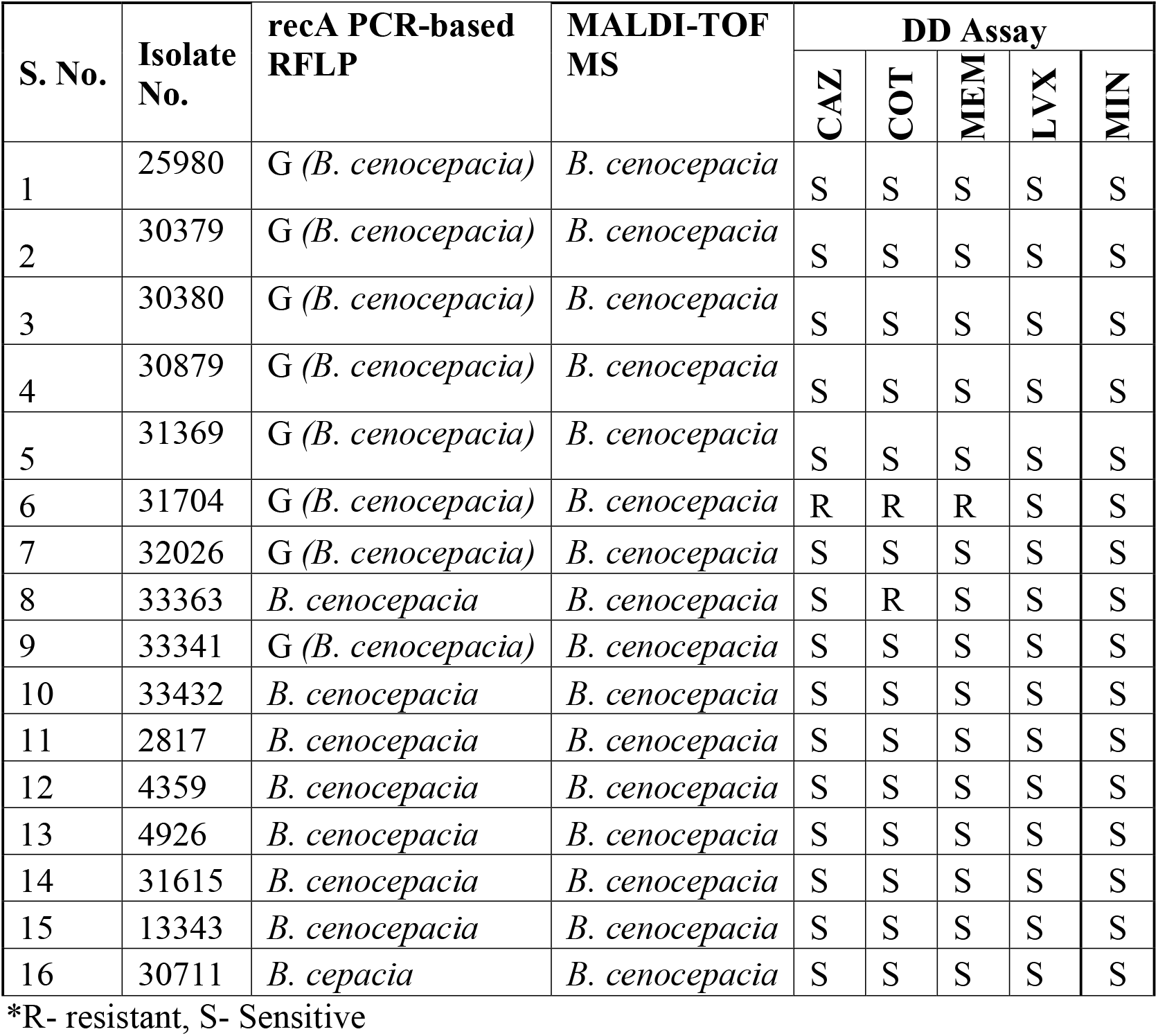
Antibiotic susceptibility test of BcenGI15 positive isolates of Bcc.

| S. No. | Isolate No. | recA PCR-based RFLP | MALDI-TOF MS | DD Assay |  |  |  |  |
| --- | --- | --- | --- | --- | --- | --- | --- | --- |
|  |  |  |  | CAZ | COT | MEM | LVX | MIN |
| 1 | 25980 | G (*B. cenocepacia*) | *B. cenocepacia* | S | S | S | S | S |
| 2 | 30379 | G (*B. cenocepacia*) | *B. cenocepacia* | S | S | S | S | S |
| 3 | 30380 | G (*B. cenocepacia*) | *B. cenocepacia* | S | S | S | S | S |
| 4 | 30879 | G (*B. cenocepacia*) | *B. cenocepacia* | S | S | S | S | S |
| 5 | 31369 | G (*B. cenocepacia*) | *B. cenocepacia* | S | S | S | S | S |
| 6 | 31704 | G (*B. cenocepacia*) | *B. cenocepacia* | R | R | R | S | S |
| 7 | 32026 | G (*B. cenocepacia*) | *B. cenocepacia* | S | S | S | S | S |
| 8 | 33363 | *B. cenocepacia* | *B. cenocepacia* | S | R | S | S | S |
| 9 | 33341 | G (*B. cenocepacia*) | *B. cenocepacia* | S | S | S | S | S |
| 10 | 33432 | *B. cenocepacia* | *B. cenocepacia* | S | S | S | S | S |
| 11 | 2817 | *B. cenocepacia* | *B. cenocepacia* | S | S | S | S | S |
| 12 | 4359 | *B. cenocepacia* | *B. cenocepacia* | S | S | S | S | S |
| 13 | 4926 | *B. cenocepacia* | *B. cenocepacia* | S | S | S | S | S |
| 14 | 31615 | *B. cenocepacia* | *B. cenocepacia* | S | S | S | S | S |
| 15 | 13343 | *B. cenocepacia* | *B. cenocepacia* | S | S | S | S | S |
| 16 | 30711 | *B. cepacia* | *B. cenocepacia* | S | S | S | S | S |

\*R- resistant, S- Sensitive

## DISCUSSION

Genomic islands are major genomic features contributing to the success of Bcc as a nosocomial pathogen, and a versatile environmental bacterium with multidrug resistance (19) (20). Fourteen GEIs namely BcenGI1-14, have already been identified in the reference genome of *B. cenocepacia* J2315 (11). In a previous study, we reported a novel 107kb genomic island namely BcenGI15 in clone ST824 associated with an outbreak in a hospital in the western part of India (16). Both the island and clone were found majorly in India. The study reported BcenGI15 with excision capability, potentially mobile with scope for further diversification. The island which incorporates the property of self-excision from its own genome and forming an extra-chromosomal circular ring, which can easily pass on via horizontal gene transfer indicated it to be an active and potentially mobile element (16). Since PGIMER (Postgraduate Institute of Medical Education and Research) is located in northern India and is a major tertiary care hospital catering to several major states of the region, screening for the BcenGI15 may be a good starting point for such study. However, the BcenGI15 harboring isolates belong to a different ST in PGIMER that is different from the ST of isolates in Mumbai hospital. The probable reasons for difference in clones may be due to unique geographic changes, horizontal gene transfers, and selection of clones in conditions prevalent there. Even within India, which has the highest patient density and heavy antibiotics usage, along with suitable tropical climatic conditions, there may be more such genomic islands yet to be identified. Hence, large-scale countrywide population genetic and genomic studies are need of the hour both in adult and pediatric populations in hospital settings.

In an earlier outbreak study, the complete sequence of BcenGI15 revealed that it harbors pathogenicity-related genes, which help bacteria survive by forming biofilms, invasion, intracellular survival, and cytotoxicity (21). In this context, further genetic, cellular, and functional studies are required. Additionally, studies are required to study role of island in different genetic background. Since the BcenGI15 is chromosomal borne, there is scope for further diversification through acquisition of more genes. Hence, there is a need to compare the BcenGI15 sequence in all the positive isolates and see variation in the island itself. Further, we need to look into the excising frequency in the different backgrounds, and more importantly, it would provide evidence for its mobilization via conjugation. Hence, studies are required to crack its role in virulence, fitness, or environment seen in hospital settings with heavy antimicrobials usage.

## CONCLUSION

Bcc is a low virulence organism having the capability of causing life-threatening infections. A proper identification of this organism is necessary to curb the dangerous outcomes due to this organism. Though comparatively resistant to various antimicrobials, this organism can be managed if a timely diagnosis with automated methods could be made out accurately. Further an insight into the novel pathogenic island possessed by this organism is an evidence that the organism is evolving rapidly in order to survive in environment leading to a rapid increase in the diseases caused by this previously known nosocomial pathogen. We concluded that presence of BcenGI15 in two different STs (ST824, ST839) from hospitals in north, west part of India that suggests probable movement, and selection for this element in Indian population of Bcc isolates. This study will probably be of immense help in understanding the mechanism and reason of nosocomial infections due to Bcc and the probable cause of emergence of an era of infection by a previously silent microbe now capable of causing havoc in the patients.

## Data Availability

All data produced in the present work are contained in manuscript

## ETHICS STATEMENT

Institutional Ethics Committee, Postgraduate Institute of Medical Education and Research (PGIMER), Chandigarh (11/6091), approved permission.

## AUTHORS AND CONTRIBUTIONS

T. S. was involved in multilocus sequence typing, analysis, and interpretation. S. K. carried out RFLP/recA-PCR, biochemical test and antibiotic susceptibility testing. R. K. revived the glycerol stocks, performed identification of isolates by MALDI-TOF and antibiotic susceptibility tests. P. P. P. designed primers for island screening. P. B. P and V. G. was involved in funding acquisition and conceptualization of the study. T. S. and C. S. drafted the manuscript; P. B. P., L. S. and V. G. revised the manuscript.

## ACKNOWLEDGMENTS

This work is supported by a project OLP148 entitled ‘High throughput and integrative genomic approaches to understand adaptation of probiotic and pathogenic bacterium’ granted to P.B.P. T.S. acknowledges senior research fellowship (SRF) from CSIR.

